# Translational Modeling Identifies Synergy between Nanoparticle-Delivered miRNA-22 and Standard-of-Care Drugs in Triple Negative Breast Cancer

**DOI:** 10.1101/2021.10.19.21265154

**Authors:** Prashant Dogra, Javier Ruiz Ramírez, Joseph D. Butner, Maria J. Peláez, Caroline Chung, Anupama Hooda-Nehra, Renata Pasqualini, Wadih Arap, Vittorio Cristini, George A. Calin, Bulent Ozpolat, Zhihui Wang

**Affiliations:** Mathematics in Medicine Program, Houston Methodist Research Institute, Houston, TX 77030, USA; Department of Physiology and Biophysics, Weill Cornell Medical College, New York, NY 10065, USA; Department of Radiation Oncology, The University of Texas MD Anderson Cancer Center, Houston, TX 77030, USA; Rutgers Cancer Institute of New Jersey, Newark, NJ 07101, USA; Department of Medicine, Division of Hematology/Oncology, Rutgers New Jersey Medical School, Newark, NJ 07103, USA; Department of Radiation Oncology, Division of Cancer Biology, Rutgers New Jersey Medical School, Newark, NJ 07103, USA; Department of Imaging Physics, The University of Texas M.D. Anderson Cancer Center, Houston, TX 77230, USA; Physiology, Biophysics, and Systems Biology Program, Graduate School of Medical Sciences, Weill Cornell Medicine, New York, NY 10065, USA; Department of Translational Molecular Pathology, The University of Texas MD Anderson Cancer Center, Houston, TX 77030, USA; Department of Experimental Therapeutics, The University of Texas MD Anderson Cancer Center, Houston, TX 77030, USA

**Author notes:** Invited Contribution. Correspondence should be addressed to: **Zhihui Wang, Ph.D.**, Associate Professor, Mathematics in Medicine Program, The Houston Methodist Research Institute.

**Keywords:** Cancer treatment, mathematical modeling, microRNA, pharmacokinetics and pharmacodynamics, precision medicine, tumor-immune interaction, allometry

## Abstract

The downregulation of miRNA-22 in triple negative breast cancer (TNBC) is associated with upregulation of eukaryotic elongation 2 factor kinase (eEF2K) protein, which regulates tumor growth, chemoresistance, and tumor immunosurveillance. Moreover, exogenous administration of miRNA-22, loaded in nanoparticles to prevent degradation and improve tumor delivery (termed miRNA-22 nanotherapy), to suppress eEF2K production has shown potential as an investigational therapeutic agent *in vivo*. To evaluate the translational potential of miRNA-22 nanotherapy, we developed a multiscale mechanistic model, calibrated to published *in vivo* data and extrapolated to the human scale, to describe and quantify the pharmacokinetics and pharmacodynamics of miRNA-22 in virtual patient populations. Our analysis revealed the dose-response relationship, suggested optimal treatment frequency for miRNA-22 nanotherapy, and highlighted key determinants of therapy response, from which combination with immune checkpoint inhibitors was identified as a candidate strategy for improving treatment outcomes. More importantly, drug synergy was identified between miRNA-22 and standard-of-care drugs for TNBC, providing a basis for rational therapeutic combinations for improved response.

## Introduction

Triple negative breast cancer (TNBC) accounts for up to 10-12% of all breast cancer cases, and has a 5-year survival that is 8-16% lower than the hormone-receptor positive (HR+) disease subtype (1). Mechanisms to overcome the aggressiveness, histopathological heterogeneity, and prevalence of TNBC in younger women represent major unmet needs in contemporary cancer medicine (2). The severity of TNBC is further aggravated due to the lack of broadly-applicable targeted therapies, and by a high-rate of early metastases to the central nervous system and lungs (3). PARP inhibitors, such as olaparib and talazoparib, have been approved for TNBC with germline BRCA1 or BRCA2 gene mutations, but these are only reported in 15.4% of cases (4). Despite recent advances in developing therapeutics for treating TNBC such as immune checkpoint inhibitor immuotherapy (5), chemotherapy remains the most common recommended systemic regimens for TNBC, even though rapid development of chemoresistance is common (6).

An emerging body of evidence has supported key functional roles of microRNAs (miRNAs) in sustaining tumor proliferation, resisting growth inhibitors and cell death, inducing tumor invasion and metastasis, and promoting angiogenesis (7), suggesting that miRNAs may function as a valuable oncologic therapy target (8-10). Amongst the multitude of miRNAs, miRNA-22 (a chain of non-coding RNA consisting of 22 nucleotides) has been found to play a critical role in cancer initiation and progression processes (11, 12). Indeed, miRNA-22 has been extensively studied as a regulator of tumor suppressor genes like p53 (13) and as a repressor of the oncogene c-Myc (14) in many different cancers, including TNBC (12), hormone-dependent breast cancer (15), and colon cancer (16), and for its roles in metastasis suppression in breast and ovarian cancer (17), in the sensitization of esophageal carcinoma to γ-ray radiation (18). miRNA-22 has been shown to be downregulated in TNBC, which reduces its inhibitory control over the eukaryotic elongation 2 factor kinase (eEF2K), a tumor growth-promoting and chemoresistance-inducing protein (12, 19). Importantly, eEF2K was also found to enhance the expression of PD-L1, and is thus implicated for its role in blocking tumor immunosurveillance (20). Inhibiting these tumorigenic effects of eEF2K via exogenous administration of miRNA-22 represents a potential therapeutic approach to improve the response of TNBC to chemotherapy and/or immunotherapy with immune checkpoint inhibitors. However, naked miRNA has a short half-life due to its vulnerability to plasma ribonucleases, shows limited tumor penetration and cellular uptake due to its negative charge, and has off-target effects due to non-specific delivery (21). To overcome these shortcomings, nanoparticle (NP)-based drug delivery systems are being studied to improve the delivery of miRNAs to tumor cells (21). While the application of nanomaterials in cancer has been promising in improving tumor imaging and drug delivery (4, 22-27), challenges associated with low tumor deliverability due to off-target accumulation and limited tumor penetration continue to limit their clinical success (28). To this end, mechanistic mathematical modeling can be a valuable in-silico tool to help overcome this challenge, by furthering our understanding of NP-mediated miRNA-22 delivery in TNBC *in vivo*.

Mathematical modeling has been used to investigate the mechanisms relevant to tumor response to miRNA-based treatment. For instance, a system of ordinary differential equations (ODEs) with a delay term has been used to study feedback loops between the oncogenes Myc, EF2, and miRNA-17-92 (29). This model was subsequently expanded by integrating nine different mechanisms to evaluate how miRNAs regulate translation (30), and to study how the inactivation of a transcription factor is involved in cardiac dysfunction and cancer (31). In another notable study (32), an energy availability pathway involving miRNA-451 was analyzed in order to elucidate the difference between invasion and proliferation regimes in cancer cells, which was accomplished by combining a pair of ODEs governing the miRNA and glucose concentration with a system of partial differential equations (PDEs) employing transport mechanisms, such as diffusion, chemotaxis, and haptotaxis. Additionally, a signaling pathway relating miRNA-21, miRNA-155, and miRNA-205 to the proliferation and apoptosis of non-small-cell lung cancer cells has been examined with a series of modeling studies (33, 34). A noteworthy feature of the mathematical approach in (34) is the inclusion of a directional migration term, which takes into account the competition for available space between cells under the assumption of a logistic growth rate for cancer cells.

While previous modeling works focused on the molecular interactions of miRNAs associated with their therapeutic outcome, they lacked the inclusion of a viable drug delivery system and the related pharmacokinetics and drug delivery mechanisms required to assess the feasibility of miRNAs as a systemically-deliverable therapy for cancer. Therefore, to support the development of miRNA-22 as a viable therapy for TNBC, here we present a mechanistic mathematical model, formulated as a system of ODEs, to describe the tumoral delivery of systemically administered miRNA-22-loaded NPs and the pharmacodynamics of miRNA-22 in the context of TNBC growth. Our multiscale model incorporates processes pertinent to systemic NP pharmacokinetics, intratumoral transport of NPs, and the known molecular interactions of miRNA-22 with its associated oncogenes to predict TNBC growth dynamics. Following model calibration with *in vivo* data that was then allometrically scaled to humans, we simulated clinically relevant treatment of TNBC with miRNA-22 to obtain the dose-response relationship at the individual and population scales, thus helping to reveal the optimal dose and frequency of treatment for each individual “virtual” patient. Local and global sensitivity analyses of key model parameters revealed the importance of molecular interactions, tumor vascularization, miRNA-22 potency, NP characteristics, and immune checkpoint effects of anti-PD-L1 in governing the outcome of miRNA-22 therapy, thus highlighting some of the key determinants of treatment outome and suggesting the potential benefit of combination with immune checkpoint inhibitors. Drug synergy was identified to occur between miRNA-22 and standard-of-care therapies (including both chemotherapy and immunotherapy) studied in this work. As a result, our mechanistic model may serve as a useful computational means to help design and optimize a therapeutic framework for future clinical trials of miRNA-22.

## Methods

### Mathematical model development

We present a multiscale mechanistic model to simulate the *in vivo* and translational pharmacokinetics (PK) and pharmacodynamics (PD) of NP-mediated miRNA-22 therapy in TNBC, alone or in combination with chemotherapy or immunotherapy (here collectively referred to as *agents*), and thus investigate the factors governing the delivery of agents and their therapeutic efficacy. The model consists of two main compartments (**Figure 1**), represented by the *plasma* and *tumor*, where the latter is sub-compartmentalized into *vascular, interstitial, cellular membrane*, and *cytosolic space*. After injection into the plasma compartment, agents are cleared through various physiological processes, which (along with the volume of distribution of agents) governs their systemic (i.e., plasma) pharmacokinetics. From the plasma compartment, bi-directional, perfusion-mediated delivery characterizes the transport of agents into the tumor vasculature, from where the extravasation of agents across the permeable tumor vasculature introduces them into the tumor interstitium. Once in the interstitium, given the absence of advection due to high interstitial fluid pressure (35, 36), the agents undergo diffusion to reach the interstitium-cell membrane interface, from where based on the type of agent, specific biophysical processes occur to ensure delivery to the target site to invoke the pharmacodynamic effects of the agents. For instance, NPs undergo endocytosis into cancer cells where they release miRNA-22 in cancer cell cytosol, whereas free drugs simply diffuse into the cytosol and antibodies bind to their corresponding cell surface receptors (e.g. PD-L1 in the current context).

**Figure 1.**
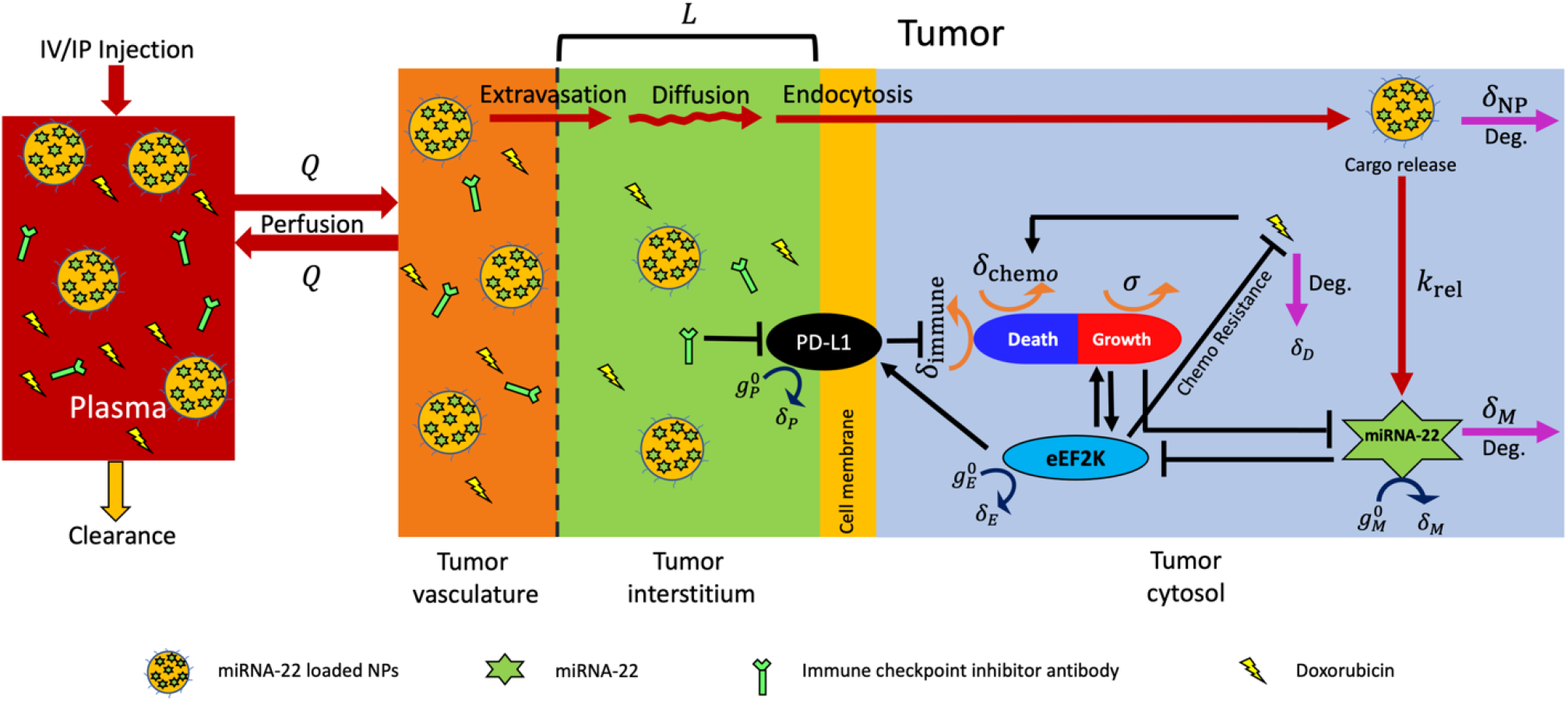
Multiscale mechanistic model. Model schematic shows key system interactions and variables. The plasma compartment is connected to the tumor compartment, with the latter sub-compartmentalized into vascular, interstitial, and cytosolic compartments. Key transport processes responsible for drug delivery to the tumor cytosol include perfusion, extravasation across tumor vasculature, diffusion across tumor interstitium, and endocytosis. While target receptors of immune checkpoint inhibitors are on the cell surface, the other agents including miRNA-22 and chemotherapeutics act intracellularly. Key signaling pathways relevant to miRNA-22 included in the model are shown in the cytosolic sub-compartment, including eEF2K induced tumor growth and PD-L1 production, miRNA-22 induced inhibition of eEF2K production, suppression of tumor antigenicity by checkpoint PD-L1, eEF2K induced chemoresistance, and induction of tumor death by chemotherapeutic agents.

Following delivery of agents to the target site, the pharmacodynamic component of the model is then engaged such that miRNA-22 acts by inhibiting the production of eEF2K protein, leading to inhibition of tumor growth. Also, because there is growing evidence in the literature that eEF2K induces the production of PD-L1 (20), this pathway was incorporated in the model to explore the engagement of immune checkpoints by eEF2K for tumor survival. The reference chemotherapeutic (i.e., doxorubicin) acts by inducing apoptotic cell death, whereas anti-PD-L1 antibodies act by inhibiting the tumor protective effects of PD-L1. Note that the mechanism of action of anti-PD-L1 antibodies modeled here includes the degradation of PD-L1 protein. Simulations and analysis of the model will help to provide insights into the systemic and tumoral pharmacokinetics of therapeutic agents and nanoparticles, along with the effects on tumor progression. We then use the model as an *in silico* tool to simulate virtual clinical trials in order to explore the effects of patient variability and other system parameters on treatment outcomes with mono- or combination therapies.

The various transport and pharmacological processes described above and shown in **Figure 1** have been formulated into a system of ordinary differential equations (ODEs; Eqs. 1–17) to obtain the temporal evolution of model behaviors of interest, including tumor growth. Equations pertaining to various compartments and biological processes are described below:

*Equation for NP mass kinetics in plasma* (*N*_P_(*t*)):

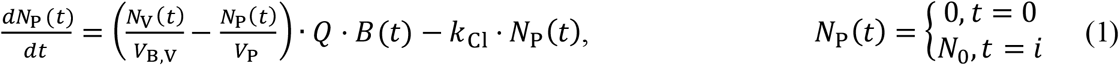

where *V*_B,V_ (= *f*_*v*_ *\* B*(*t*)) and *V*_P_ are volumes of tumor vasculature and plasma compartments, respectively; *f*_*v*_ is the vascular volume fraction of the tumor; *B*(*t*) represents tumor volume; *Q* represents plasma flow rate; *k*_cl_ represents systemic clearance of NPs; *N*_0_ is the injected dose of NPs; and *i* represents the injection times (in weeks) post inoculation of tumor in mice (*i* = 2, 3, 4, 5). Note that *Q* (units, mL · mL^−1^ · wk^−1^) obeys the following empirical relationship with tumor volume (*B*(*t*)): *Q* = 2843 · *e*^−0.65·*B*(*t*)^, obtained by fitting a monoexponential function to data from literature (37). Note that for human simulations, *Q* was assumed to be 1512 mL · mL^−1^ wk^−1^ irrespective of tumor size (38). Given that tissue density is ∼1 g · mL^−1^, tumor blood flow rate is provided in the units of mL · mL^−1^ · wk^−1^ in our work, which is numerically equivalent to mL · g^−1^ · wk^−1^ as typically used in the literature. In addition, NP clearance in mice *k*_cl_ (units, wk^−1^) varies empirically with NP diameter (*ϕ*_NP_ ; units, cm) as: 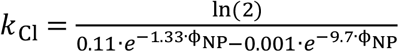, obtained by fitting the plasma half-life data of quantum dots of varying sizes from the literature (39, 40). For human simulations, the value of *k*_Cl_ was allometrically scaled (see next Section).

*Equation for NP mass kinetics in tumor vasculature* (*N*_V_(*t*)):

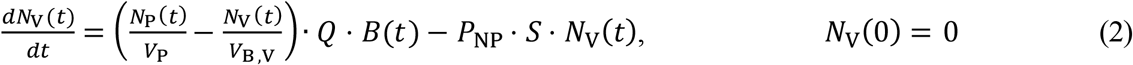

where *P*_NP_ indicates NP permeability across tumor vasculature and *S* is the tumor vascular surface area per unit tumor volume (units, cm^2^/cm^3^). In vivo, *S* relates to tumor volume *B*(*t*) as *S* = 0.26 · *e* ^−4.5·*B*(*t*)^ + 138 · *e*^−0.04·*B*(*t*)^, obtained empirically from the literature (41), and *P*_NP_ (units, cm · wk^−1^) is a function of tumor vascular porosity and the ratio of NP size (*ϕ*_NP_) to tumor vascular pore size (*ϕ*_pore_; units, cm) (42). For human simulations, *S* was fixed at 135 cm^2^⁄cm^3^ irrespective of tumor size (41).

*Equation for NP mass kinetics in tumor interstitium* (*N*_V_(*t*)):

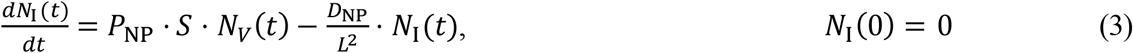

where *D*_NP_ is the diffusivity of NPs in tumor interstitium, and *L* is the characteristic interstitial distance between tumor vessels and cancerous cells, referred to herein as the intercapillary length in the tumor.

*Equation for NP mass kinetics in cancer cell membrane* (*N*_M_(*t*)):

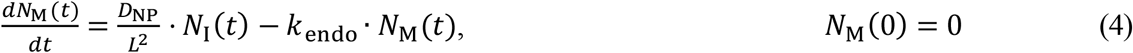

where *k*_endo_ (units, wk^−1^) is the rate of endocytosis of NPs into tumor cells, and may be obtained by equating work done by the membrane motor proteins against surface tension of cell membrane (43).

*Equation for NP mass kinetics in cancer cell cytosol* (*N*_*C*_(*t*)):

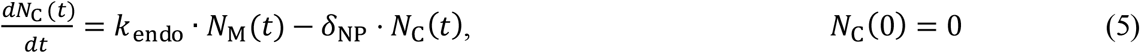

where *δ*_NP_ is NP degradation rate.

*Equation for miRNA concentration kinetics in cancer cell cytosol* (*C*_M_(*t*)):

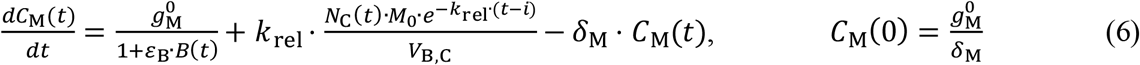

where 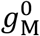 is the intrinsic production rate of miRNA-22 in the tumor cytosol; *ε*_B_ is the efficiency of tumor on inhibiting miRNA-22 production; *M*_0_ is the mass of miRNAs loaded in a single NP; and *V*_B,C_ is the cytosolic volume of tumor (= *f*_*c*_ *\* f*_cy_ *\* B*(*t*)), where *f*_*c*_ (= 0.4) is the cancer cell volume fraction of a tumor (44) and *f*_cy_ (= 0.4) is the cytoplasmic volume fraction of a cancer cell (45). *k*_rel_ is release rate of miRNAs from the endocytosed NP; and *δ*_M_ is the degradation rate of miRNAs. Note that the initial condition *C*_M_(0) is estimated at the trivial steady state of the system when *B*(0) = 0 and no exogenous administration of miRNAs has occured, such that 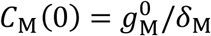.

*Equation for anti-PD-L1 antibody concentration kinetics in plasma* (*C*_Ab,P_(*t*)):

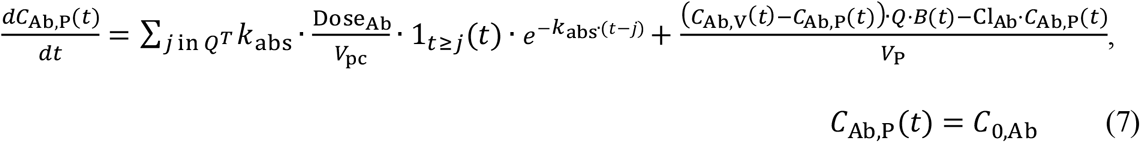

where Dose_Ab_ represents the dose of antibodies; *k*_abs_ is the absorption rate constant of antibodies from the peritoneum into plasma after intraperitoneal (IP) injection; *V*_pc_ is the volume of peritoneal fluid in female mice (0.1 mL) (46); Cl_Ab_ is the systemic clearance of antibodies; and *j* represents the injection times (in weeks) post inoculation of tumor in mice defined in the set *Q*^*T*^ (= 1, 1.71, 2.43, 3.14, 3.86, 4.57). Note that in mouse experiments, antibodies were given IP; therefore, the initial plasma concentration of antibodies *C*_0,Ab_ is zero. However, for human simulations, immunotherapy was administered as an intravenous (IV) bolus injection, therefore the initial plasma concentration of antibodies *C*_0,Ab_ is non-zero and is calculated based on the initial dose (Dose_Ab_) and systemic volume of distribution (*V*_D,Ab_) of the given antibody. As a result, the first term of Eq. 7 that accounts for systemic absorption of the drug from the peritoneum is removed during human simulations.

*Equation for anti-PD-L1 antibody concentration kinetics in tumor vasculature* (*C*_Ab,v_(*t*)):

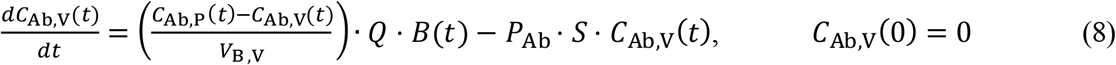

where *P*_Ab_ indicates antibody permeability across tumor vasculature.

*Equation for anti-PD-L1 antibody concentration kinetics in tumor interstitium* (*C*_Ab,I_(*t*)):

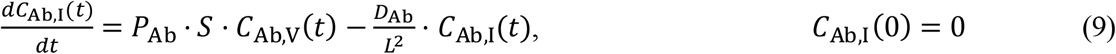

where *D*_Ab_ is the diffusivity of antibodies in tumor interstitum.

*Equation for anti-PD-L1 antibody concentration kinetics in cancer cell membrane* (*C*_Ab,M_(*t*)):

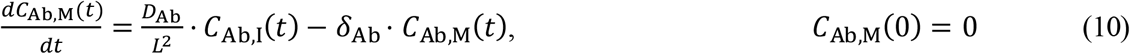

where, *δ*_Ab_ is the degradation rate of antibodies.

*Equation for doxorubicin concentration in plasma* (*C*_D,P_(*t*)):

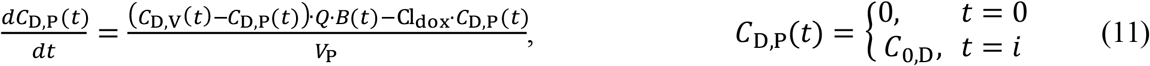

where Cl_dox_ is the plasma clearance of doxorubicin; *C*_0,D_ is the initial concentration of doxorubicin calculated based on the injected dose of 4mg/kg and the given volume of distribution (**Table 2**); *i* represents the injection times (in weeks) post inoculation of tumor in mice (*i* = 1, 2, 3).

*Equation for doxorubicin concentration in tumor vasculature* (*C*_D,V_(*t*)):

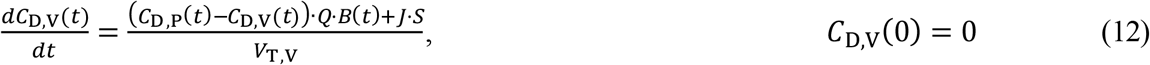

where *J* is the diffusive flux of doxorubicin across the tumor vasculature into tumor interstitium, and is given by 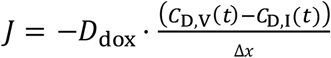. Here,*D* _*dox*_ is the diffusivity of doxorubicin in tumor interstitium and Δ*x* is the thickness of blood capillary wall (5 μm) (47).

*Equation for doxorubicin concentration in tumor interstitium* (*C*_D,I_(*t*)):

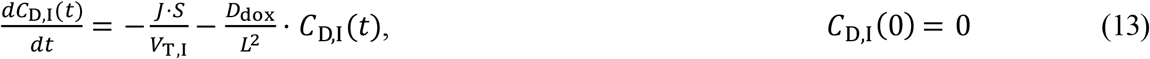

*Equation for doxorubicin concentration in tumor cytosolic space* (*C*_D,C_(*t*)):

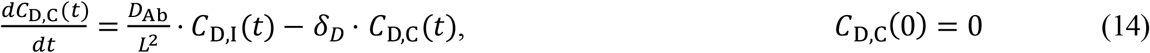

where *δ*_D_ is the degradation rate of doxorubicin in the cytosolic space.

*Equation for eEF2K concentration kinetics in cancer cell cytosol* (*C*_E_(*t*)):

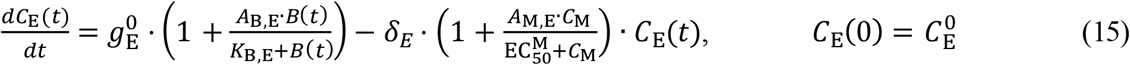

where 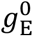 is the basal production rate of eEF2K protein in tumor cytosol; *A*_B,E_ is the stimulation factor of tumor effects on eEF2K production; *K*_B,E_ is the Michaelis-Menten constant for tumor effects on eEF2K production; *δ*_E_ is the degradation rate of eEF2K protein; *A*_M,E_ is the stimulation factor of miRNA-22 effects on eEF2K protein degradation; 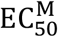 is the half-maximal effective concentration of miRNA-22 for its effect on eEF2K protein degradation; and 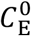 is the intial concentration of eEF2K protein.

*Equation for PD-L1 concentration kinetics in cancer cell membrane* (*C*_P_(*t*)):

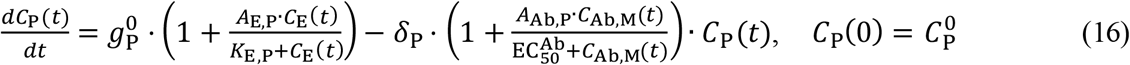

where 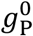 is the basal production rate of PD-L1 protein in tumor cytosol; *A*_E,P_ is the stimulation factor of eEF2K effects on PD-L1 production; *K*_E,P_ is the Michaelis-Menten constant for eEF2K effects on PD-L1 production; *δ*_P_ is the degradation rate of PD-L1 protein; *A*_Ab,P_ is the stimulation factor of anti-PD-L1 antibody effects on PD-L1 degradation; 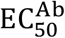 is the half-maximal effective concentration of anti-PD-L1 antibody for its effect on PD-L1 protein degradation; and 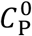 is the intial concentration of PD-L1 protein.

*Equation for tumor volume kinetics* (*B*(*t*)):

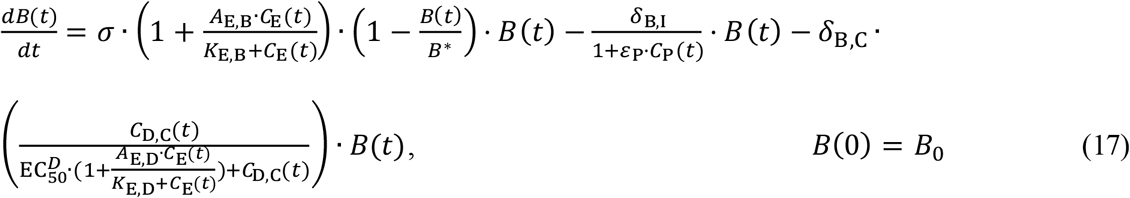

where *σ* is the tumor growth rate constant; *A*_E,B_ is the stimulation factor representing eEF2K effects on tumor growth; *K*_E,B_ is the Michaelis-Menten constant for eEF2K effects on tumor growth; *B*^*\**^ is the tumor carrying capacity; *δ*_B,I_ is the death rate of tumors induced by normal immune system functionality (without drug intervention); *ε*_P_ is the efficiency of PD-L1 protein in inhibiting immune-induced tumor death; *δ*_B,C_ is the death rate of tumors induced by doxorubicin; EC_50_^*D*^ is the half-maximal effective concentration of doxorubicin for its effect on tumor death; *A*_E,D_ is the stimulation factor for eEF2K effects in inducing chemoresistance; *K*_E,D_ is the Michaelis-Menten constant for eEF2K effects in inducing chemoresistance; and *B*_0_ is the size of inoculated tumor, i.e., initial condition (equal to a single cell volume for human simulations).

The model was solved numerically as an initial value problem in MATLAB R2018a using the built - in function *ode15s*, and fit to the *in vivo* data from literature (12, 48-50) using the built-in function *lsqcurvefit*. Correlation analysis was then performed between model fits and experimental data to assess the goodness of fit.

### Allometric scaling

For human simulations, the rate constants *k*_Cl_, *σ, δ*_B,I_, and *δ*_B,C_ were allometrically scaled from values determined for mice, based on body weights and the standard allometric exponent for rate constants, i.e., –0.25 (51), such that the value of parameter *i* for humans 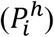 was: 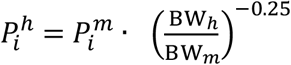, where 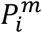 is the value of parameter *i* for mice, and BW_*h*_ and BW_*m*_ are the body weights assumed for humans (70 kg) and mice (0.02 kg), respectively. We note that a different scaling exponent was used in the above formula for a subset of parameters, as based on published reports, these were: dose and clearance calculations (exponent = 0.75) and volume of distribution calculations (exponent = 1.0) (52, 53).

### Treatment response evaluation

The model was used to study the effect of miRNA-22 nanotherapy, alone or in combination with doxorubicin and/or Atezolizumab, in virtual patients. The effect of therapy on tumor shrinkage was quantified by a metric defined as percent tumor growth inhibition (%TGI), such that %TGI = (1 −*B*_treated_/*B*_control_) · 100, where *B*_treated_ and *B*_control_ represent treatment and control tumor volumes at the end of 104 weeks post tumor inception with a single cell. Note that, in the treatment scenario, therapy was initiated 80 weeks post tumor inception, such that treatment was given over 24 weeks (∼6 months). To evaluate tumor response at a population level, we employed a scale analogous to RECIST 1.1 (50), such that treatment response was classified as *progressive disease* (TGI ≤ 0%), *stable disease* (0% < TGI ≤ 10%), *intermediate response* (10% < TGI ≤ 30%), *partial response* (30% < TGI ≤ 50%), and *major response* (TGI > 50%).

### Parameter sensitivity analysis

To investigate the importance of various model parameters in causing tumor shrinkage in patients undergoing treatment with a weekly dose of 0.026 mg/kg miRNA-22 (loaded in NPs), we performed local (LSA) and global (GSA) sensitivity analyses (42, 54-57) by perturbing the parameters of interest (highlighted by a dagger in **Tables 1, 2**) over a range of 0.2× to 5× of their corresponding baseline values.

**Table 1.** List of biological parameters and initial conditions.

| Parameter | Description | Units | Value | Ref. |
| --- | --- | --- | --- | --- |
| **eEF2K-related parameters** |  |  |  |  |
| $A_{B,E}^{\dagger}$ | Stimulation factor for tumor effects on eEF2K production | — | 11.9 | Est |
| $A_{M,E}^{\dagger}$ | Stimulation factor for miRNA-22 effects on eEF2K degradation | — | 10.52 | Est |
| $K_{B,E}^{\dagger}$ | Michaelis-Menten constant for tumor effects on eEF2K production | cm<sup>3</sup> | 16.03 | Est |
| $\delta_E$ | Decay rate of eEF2K | wk<sup>-1</sup> | 60.48 | (34) |
| $g_E^{0\dagger}$ | Basal production rate of eEF2K protein | wk<sup>-1</sup> | 36.3 | Est |
| $C_E^0$ | eEF2K initial condition | — | 0.58 | Est |
| **PD-L1-related parameters** |  |  |  |  |
| $A_{E,P}^{\dagger}$ | Stimulation factor for eEF2K effects on PD-L1 production | — | 3.32 | Est |
| $A_{Ab,P}$ | Stimulation factor for anti-PD-L1 antibody effects on PD-L1 degradation | — | 1.79 | Est |
| $K_{E,P}^{\dagger}$ | Michaelis-Menten constant for eEF2K effects on PD-L1 production | — | 8.1 | Est |
| $\delta_P$ | Decay rate of PD-L1 | wk<sup>-1</sup> | 60.48 | (34) |
| $g_P^{0\dagger}$ | Basal production rate of PD-L1 protein | wk<sup>-1</sup> | 10.44 | Est |
| $C_P^0$ | PD-L1 initial condition | — | 0.21 | Est |
| **Tumor-related parameters** |  |  |  |  |
| $A_{E,B}^{\dagger}$ | Stimulation factor for eEF2K effects on tumor growth | — | 4.5 | Est |
| $K_{E,B}^{\dagger}$ | Michaelis-Menten constant for eEF2K effects on tumor growth | — | 8.78 | Est |
| $A_{E,D}$ | Stimulation factor for eEF2K effects on inducing chemoresistance | — | 0.1 | Est |
| $K_{E,D}$ | Michaelis-Menten constant for eEF2K effects on inducing chemoresistance | — | 10.0 | Est |
| $\sigma^{\dagger}$ | Tumor growth rate constant | wk<sup>-1</sup> | 3.1 (miRNA-22, M)<br>3.75 (Dox, M)<br>3.13 (Atezo, M)<br>0.43 (H) | Est, Allo |
| $B^*$ | Tumor carrying capacity | cm<sup>3</sup> | 2.21 (miRNA-22, M)<br>2.5 (Dox, M)<br>2.99 (Atezo, M)<br>100 (H) | Est |
| $\delta_{B,I}^{\dagger}$ | Tumor death rate (immune-induced) | wk<sup>-1</sup> | 3.0 (M), 0.39 (H) | Est, Allo |
| $\varepsilon_P^{\dagger}$ | Efficiency of PD-L1 to inhibit immune-induced tumor death | — | 1.9 | Est |
| $\delta_{B,C}$ | Tumor death rate (chemo-induced) | wk<sup>-1</sup> | 2.46 (M), 0.3198 (H) | Est, Allo |
| $\phi_{\text{pore}}$ | Diameter of tumor vessel wall pores | nm | 1700 | (65) |
| $L$ | Intercapillary length | cm | 0.01 | (66) |
| $\eta_{B,B}$ | Dynamic viscosity of tumor blood | cP | 7.42 | (41) |
| $\eta_{B,I}$ | Dynamic viscosity of tumor interstitium | cP | 3.5 | (67) |
| $B_0$ | Tumor initial condition | cm<sup>3</sup> | 0.001 | (12) |
| $f_v$ | Tumor vascular volume fraction | — | 0.17 | (41) |
| $Q^\dagger$ | Tumor blood flow rate | $\text{mL} \cdot \text{mL}^{-1} \cdot \text{wk}^{-1}$ | $Q = 2843 \cdot e^{-0.65 \cdot B(t)}$<br>(M), 1512 (H) | (37, 38) |
| $S^\dagger$ | Tumor microvascular surface area | $\text{cm}^2/\text{cm}^3$ | $S = 0.26 \cdot e^{-4.5 \cdot B(t)} + 138 \cdot e^{-0.04 \cdot B(t)}$ (M),<br>135 (H) | (41) |
| **Systemic circulation-related parameters** |  |  |  |  |
| $V_p$ | Volume of plasma compartment | mL | 1 (M), 3000 (H) | (61) |
| $V_{pc}$ | Volume of peritoneal fluid | mL | 0.1 (M) | (46) |

Note: Mice and human specific parameters are specified by M and H in parantheses, respectively. Not specified in case of common values. <sup>†</sup>Dagger indicates patient-specific parameters perturbed for virtual clinical trial simulations. Abbreviations: Est- estimated via regression, Allo- allometrically scaled, Dox- doxorubicin, Atezo- Atezolizumab.

**Table 2.** List of therapy-related parameters and initial conditions.

| Parameter | Description | Units | Value | Ref. |
| --- | --- | --- | --- | --- |
| **NP-related parameters** |  |  |  |  |
| $\phi_{NP}$ | NP diameter | nm | 70 | (12) |
| $\delta_{NP}$ | NP degradation rate | $\text{wk}^{-1}$ | 7.7 | (68) |
| $N_0$ | Number of NPs per injection | — | $\sim 2.5\text{e}+10$ | Calc |
| **miRNA-22-related parameters** |  |  |  |  |
| $EC_{50}^M$ | $EC_{50}$ of miRNA-22 | nM | 2.34 | Est |
| $k_{rel}$ | Release rate of miRNA-22 from NPs | $\text{wk}^{-1}$ | 0.99 | Est |
| $g_M^{0\dagger}$ | Basal production rate of miRNA-22 | $\text{nM} \cdot \text{wk}^{-1}$ | 0.033 | Est |
| $\varepsilon_B^\dagger$ | Efficiency of tumor to inhibit miRNA-22 production | $\text{cm}^{-3}$ | 1 | Assu |
| $\delta_M$ | Decay rate of miRNA-22 | $\text{wk}^{-1}$ | 4.851 | (69) |
| $M_0$ | miRNA-22 initial condition | nM | 0.007 | Est |
| **Chemotherapy-related parameters (Doxorubicin)** |  |  |  |  |
| $Cl_{dox}$ | Plasma clearance of dox | $\text{mL} \cdot \text{wk}^{-1}$ | 8.4e+3 (M), 4.25e+6 (H) | (70, 71) |
| $V_{D,dox}$ | Volume of distribution of dox | mL | 734 (M), 3.65e+5 (H) | (70, 71) |
| $\delta_D$ | Degradation rate of dox | $\text{wk}^{-1}$ | 2.0 | Est |
| $EC_{50}^D$ | $EC_{50}$ of dox | nM | 25 | (70) |
| **Immunotherapy-related parameters (Atezolizumab)** |  |  |  |  |
| $\phi_{Ab}$ | Ab diameter | nm | 10 | (72) |
| $\delta_{Ab}$ | Degradation rate of Ab | $\text{wk}^{-1}$ | 1.21 | Est |
| $Cl_{Ab}$ | Plasma clearance of Ab | $\text{mL} \cdot \text{wk}^{-1}$ | 3.07 (M), 1400 (H); | Allo, (73) |
| $V_{D,Ab}$ | Volume of distribution of Ab | mL | 1.97 (M), 6900 (H); | Allo, (73) |
| $EC_{50}^{Ab}$ | $EC_{50}$ of Ab | nM | 0.0446 | (74) |
| $k_{abs}$ | Peritoneal absorption rate constant of Ab | $\text{wk}^{-1}$ | 100 | Est |

Note: <sup>†</sup>Dagger indicates patient-specific parameters perturbed for virtual clinical trial simulations. Mice and human specific parameters are specified by M and H in parantheses, respectively. Abbreviations: Est- estimated via regression, Allo- allometrically scaled., Calc- calculated from formulae, Assu- assumed, Dox- doxorubicin, Ab- antibody.

LSA involved perturbation of one model parameter at a time at 500 levels between the range of 0.2× to 5× of the baseline value while the other parameters were held constant at baseline. Each parameter was perturbed individually and %TGI was calculated to obtain the qualitative relationship between parameter factor change and %TGI. Alternatively, in GSA, all model parameters of interest were simultaneously perturbed and %TGI calculated for each simulation (i.e., for a given combination of parameter values). Note that, to comprehensively investigate the vast multiparameter space (21 parameters), yet to minimize the number of simulations, Latin hypercube sampling (LHS) (42, 54, 55) was used to obtain 10,000 combinations of parameter values, and 10 such replicates were obtained. Multivariate linear regression analysis was then performed on every replicate, and regression coefficients were determined as a quantitative measure of parameter *sensitivity index* (SI). A distribution of regression coefficients (or SI) was obtained for each parameter, and one-way ANOVA with Tukey’s test was used to rank the parameters in terms of their sensitivity, such that a higher SI represents a greater influence on model output (i.e., %TGI).

### Determination of drug synergy

The Chou-Talalay method (58) was used to identify drug synergy between miRNA-22 and its combination with standard-of-care drugs for TNBC (doxorubicin and/or Atezolizumab). Occurence of drug synergy allows the possibility of using a lower dose of the constituent drugs, which can reduce their adverse effects. The method involves determination of *combination index* (CI), such that CI < 1 is an indicator of existence of drug synergism. To calculate CI, the open source software COMPUSYN (available at https://www.combosyn.com/) was used to generate the analysis report, which has been provided in the **Supplementary Information**.

## Results and Discussion

### Model development, calibration, and baseline solution

The multiscale mechanistic model developed to study the PK-PD of NP-mediated miRNA-22 therapy in TNBC, along with other clinically approved treatment modalities, was formulated as a system of ODEs (Eqs. 1-17) and solved numerically as an initial value problem. Some model parameters were known a priori (**Tables 1, 2**), while the rest were estimated through non-linear least squares fitting of the model to published *in vivo* datasets. The selected datasets include longitudinal measurements of tumor volume in mice bearing MDA-MB-231 xenografts under control conditions, or under treatment with one of the following therapies: 0.15 mg/kg (equivalent 4 μg/mouse) IV miRNA-22-loaded NPs once a week (12), 4 mg/kg IV doxorubicin once a week (48), and 5 mg/kg IP anti-PD-L1 immunotherapy (Atezolizumab) once every 5 days (50).

We used the model to simulate the treatment protocols shown in **Figure 2**, and the numerical solutions of tumor volume kinetics were then fit simultaneously to the four datasets to estimate the unknown model parameters (given in **Tables 1, 2**). Additionally, the model solution for eEF2K protein kinetics from the miRNA-22 simulation was fit to the available data (**Figure 2a**). Model fits were in good agreement with the experimental data, as indicated by a strong Pearson correlation (**Figure S1;** *R* > 0.96, *P* < 0.0001). While the various experimental studies used above demonstrated the effects of individual therapies on TNBC progression, the model revealed additional insights into drug (and also NP) pharmacokinetics and molecular interaction dynamics leading to tumor response to the three therapies.

**Figure 2.**
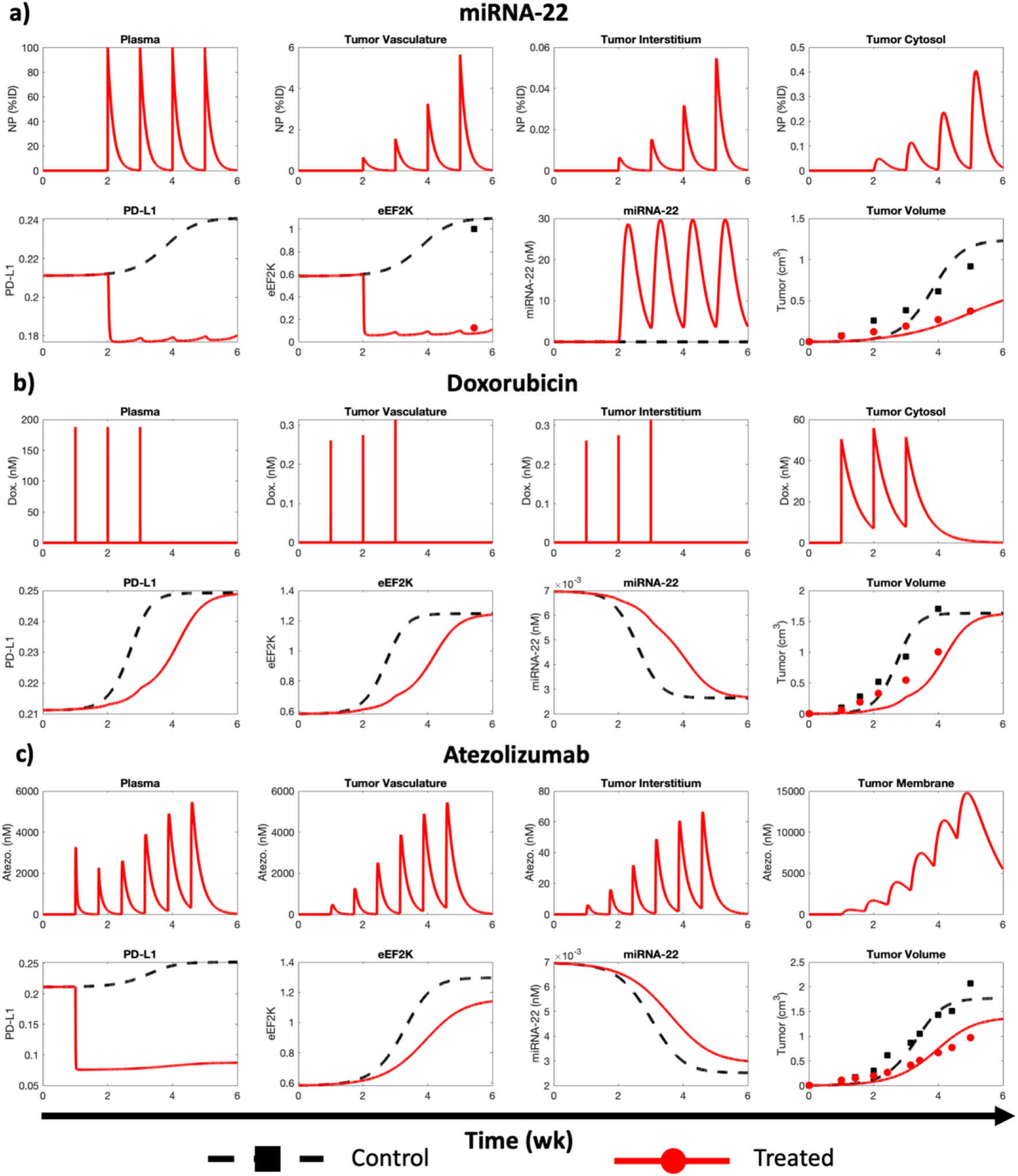
Model calibration. Numerical solution of model fit to published *in vivo* data for treatment of MDA-MB-231 tumor-bearing mice with **a**) NP-delivered miRNA-22, **b**) doxorubicin, and **c**) Atezolizumab. Markers represent experimental data. Pearson correlation analysis results goodness of fit of the model are reported in Figure S1.

As shown in **Figure 2a**, simulated NP-mediated miRNA-22 therapy involved periodic IV administration of miRNA-22-loaded NPs into the plasma compartment, from where the NPs were cleared in a size-dependent fashion characterized by *k*_cl_, and also transported to the tumor vascular sub-compartment in a perfusion-dependent manner governed by the plasma flow rate *Q*. Extravasation of NPs across the leaky tumor vasculature into the tumor interstitium, determined by the vascular permeability-surface area product (*P*_NP_ *· S*), was followed by NP size-dependent diffusion through the tumor interstitium. Remaining NPs were ultimitely endocytosed into the tumor cytosolic sub-compartment (i.e., cancer cell cytosol), followed by NP degradation and release of miRNA-22 into the cancer cell Interior. As a result, miRNA-22-induced inhibition of eEF2K production was observed relative to the control case, which reduced the downstream production of cancer cell transmembrane protein PD-L1. The overall effect of miRNA-22 therapy on alterations in protein expression manisfested as tumor growth inhibition mediated by suppressed induction of tumor growth by eEF2K and increased vulnerability to tumor immunogenicity due to depletion of the immune checkpoint PD-L1.

To explore the therapeutic combinations of miRNA-22 with FDA approved chemotherapies and immunotherapies, we calibrated the model with *in vivo* treatment of TNBC with doxorubicin (**Figure 2b**) and Atezolizumab (**Figure 2c**). In response to doxorubicin therapy (**Figure 2b**), the model showed reduction in tumor growth relative to the control case due to drug concentration-dependent increase in tumor death rate *δ*_B,C_. This is accompanied by reduced expression levels of eEF2K and PD-L1, but increased basal miRNA-22 expression level. Note that tumor growth has an inhibitory effect on miRNA-22 production (12), but stimulates eEF2K production, which tends to stimulate tumor growth in a complimentary feedback process (12, 19). We next modeled the effect of anti-PD-L1 immunotherapy (Atezolizumab) in a simplistic fashion by targeting PD-L1 degradation rate *δ*_P_, such that Atezolizumab enhances the degradation of PD-L1 in a drug concentration-dependent manner. As a result, as shown in **Figure 2c**, due to depletion of PD-L1, there is inhibition of tumor growth compared to the control. The parameters estimated as a result of the above model calibrations are given in **Tables 1, 2**.

### Model extrapolation to human scale

To study the translational value of miRNA-22 and associated potential combination therapies, we extrapolated the *in vivo* mechanistic model to human scale, either by substituting known physiological parameters with human values, or by allometric scaling of unknown parameters from mice to humans (see *Allometric Scaling* in **Methods**). In **Figure 3a**, a representative simulation of NP-mediated miRNA-22 therapy in a virtual adult patient (body weight 70 kg) is shown following once a week (QW) IV administration of 0.026 mg/kg miRNA-22 (allometrically scaled dose) for six months, starting 80 weeks after the inception of tumor with a single cell. As shown, eEF2K and PD-L1 levels are suppressed throughout the duration of treatment, thereby leading to ∼29% TGI compared to the control case. The prameters used for the representative simulation are the baseline values shown in **Tables 1, 2**.

**Figure 3.**
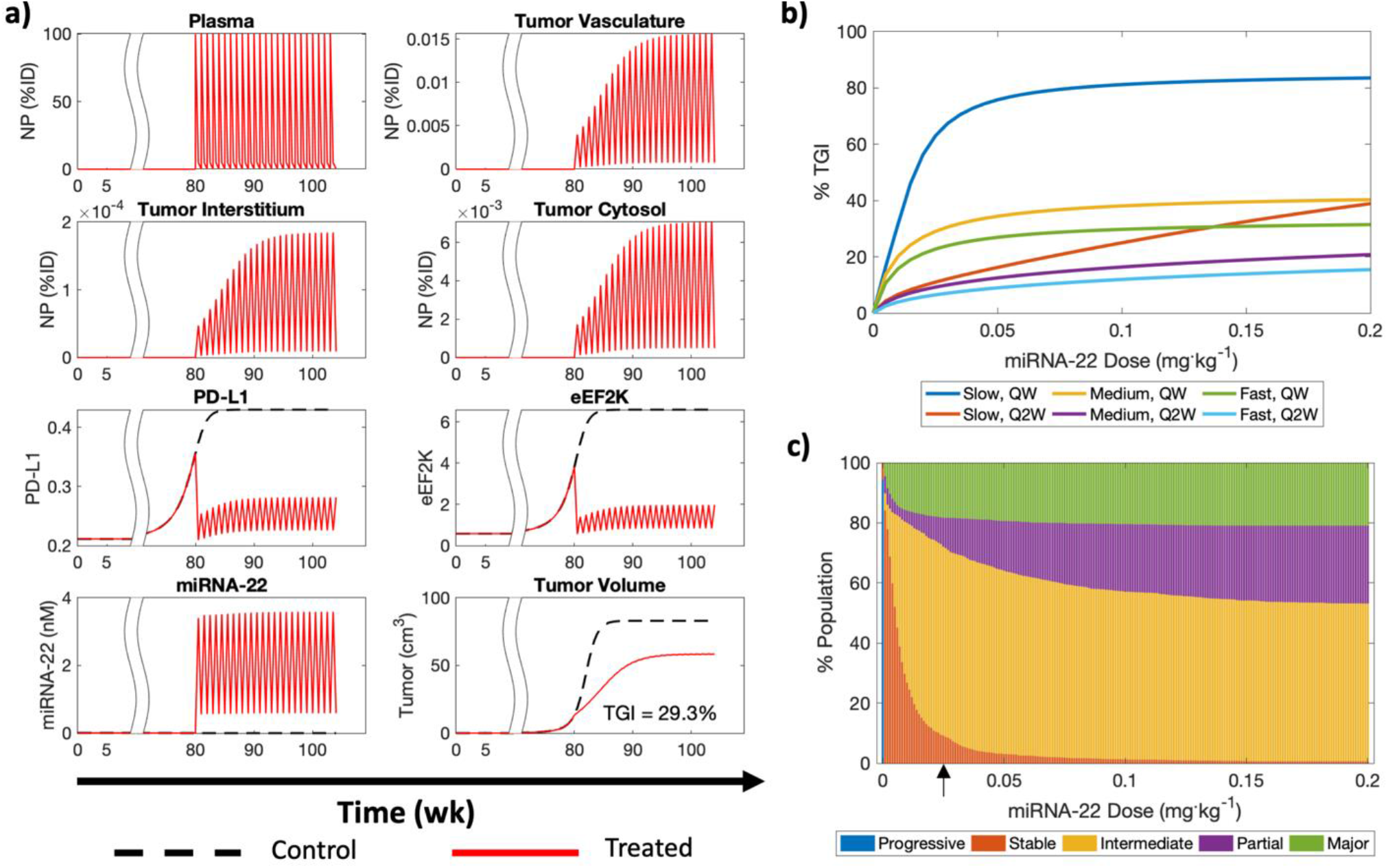
Translational PK-PD of miRNA-22. **a**) Human extrapolation of *in vivo* mechanistic model simulating treatment with once weekly dose of miRNA-22 for six months. TGI indicates percent tumor growth inhibition. **b**) Dose response curves of a virtual patient under scenarios of once weekly (QW) or once every two weeks (Q2W) dose of miRNA-22 for slow and fast growing tumors. **c**) Effects of inter-individual variability on miRNA-22 therapy outcome for different QW doses, presented on a scale analogous to RECIST 1.1. Black arrow on x-axis indicates the dose of 0.026 mg/kg used for further analysis.

### Dose-response relationship and population variability

To investigate the effect of changes in miRNA-22 doseage and treatment frequency on %TGI, dose-response curves (DRCs) was generated for a representative individual by simulating treatment with miRNA-22 nanotherapy at different doses (0-0.2 mg/kg), either once weekly (QW) or once every two weeks (Q2W) (**Figure 3b**). DRCs were also generated for slow, medium, and fast growing tumors at the two treatment frequencies. Note that the ratio of tumor immunogenicity-induced death rate (e.g., tumor death rate due to normal immune effects without drug intervention) to tumor growth rate (*δ*_B,I_/*σ*) indiciates how fast a tumor grows; e.g., within the scope of this work, the ratios of 0.99, 0.9, and 0.75 indicate slow, medium, and fast growing tumors, respectively (**Figure S3**). As shown in **Figure 3b**, theraputic response tends to saturate around a dosage of 0.05 mg/kg in these six scenarios, and even beyond 0.026 mg/kg (dose obtained through allometric scaling; see *Allometric Scaling* in **Methods**), tumors do not exhibit significant increase in %TGI, hence 0.026 mg/kg was chosen as the reference dose in humans for further investigation. As for the slow growing tumors, they show a much higher response to therapy (∼two-fold) than their rapidly proliferating counterparts. Additionally, irrespective of the rate of tumor growth, the QW protocol causes greater %TGI than Q2W.

Further, by creating a virtual population of 2,000 patients through LHS of patient-specific parameters between ±50% of their baseline values, the effects of inter-individual variability on %TGI for different doses of QW miRNA-22 were investigated and presented in a manner *analogous* to the RECIST 1.1 classification (51, 59). As shown in **Figures 3c** and **S2**, the patient population showed significant improvement in response with increasing dose up to ∼0.02 mg/kg, such that stable disease (0% < TGI ≤ 10%) cases dropped exponentially, and the population of intermediate responders (10% < TGI ≤ 30%) and major responders (> 50% TGI) grew rapidly. Also, a steady increase was observed in the population of partial responders (30% < TGI ≤ 50%). However, beyond ∼0.02 mg/kg dosage, the population of major responders quickly saturated at a value of ∼20%, while the population of partial responders increased with increasing drug dosage up to ∼0.1 mg/kg, eventually settling at ∼25%; the remaining population (∼55%) primarily consisted of intermediate responsers. Thus, these observations support our use of the allometrically calculated dose of 0.026 mg/kg; this was used as the reference value for further analysis. Note that progressive disease (≤ 0% TGI) was only seen in the no treatment scenario; this indicates that treated tumors do not grow beyond the size of the corresponding control tumors, and hence as per our definition of %TGI, ≤ 0% values are not observed under treatment. These simulations provide quantification of the variation in treatment response that can be expected from physiological variability and tumor heterogeneity on a population scale, and can thus support treatment personalization to maximize patient benefit. Note that the parameters used for virtual population generation are marked by a dagger in **Tables 1**,**2**.

### Parameter sensitivity analysis

For a more complete understanding of the effects of both patient-specific and treatment-related parameters on %TGI following 0.026 mg/kg QW dose of miRNA-22 nanotherapy for six months, starting 80 weeks post initiation of tumor, we performed local (LSA) and global (GSA) sensitivity analyses by perturbing parameters over a range of 0.2× to 5× of the baseline values (42). As shown in **Figure 4a**, GSA ranked the 21 model parameters into eight categories based on their sensitivity indices (using one-way ANOVA and Tukey’s test), out of which we discuss the top five ranking parameter brackets below.

**Figure 4.**
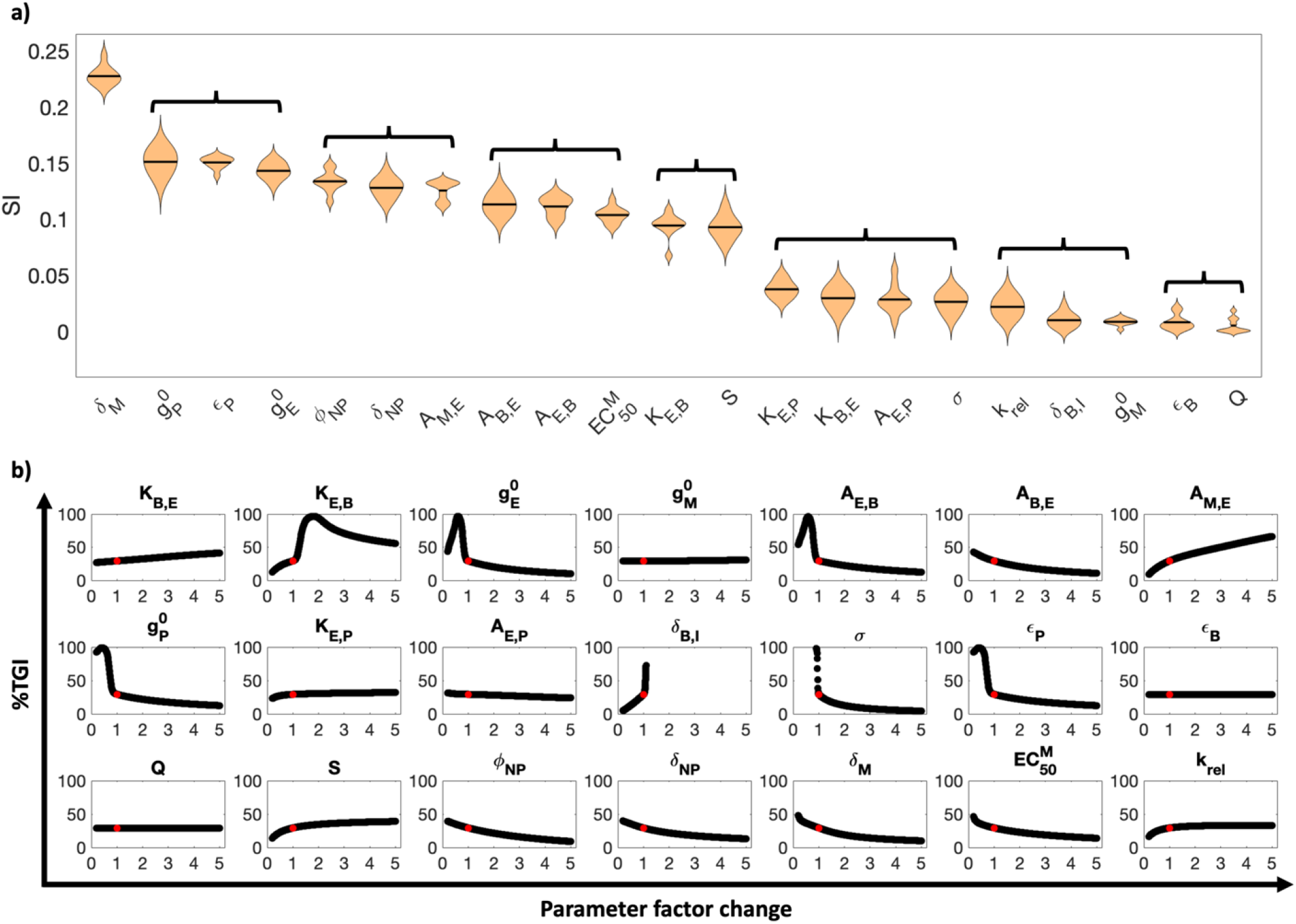
Parameter sensitivity analysis. **a**) Violin plot showing results of global sensitivity analysis such that parameters are plotted in a descending order of sensitivity from left to right. SI denotes sensitivity index. Parameters are bracketed based on their ranking obtained from Tukey’s test. **b**) Efects of individual parameters on %TGI is shown via local sensitivity analysis. Note that for both analyses, parameters were perturbed over a range of 0.2x to 5x of the baseline value. Red dot in each curve indicates the %TGI value corresponding to the baseline parameter values.

First, as shown in **Figure 4a**, miRNA-22 degradation rate (*δ*_M_) stands out for its influence on %TGI, indicating that the stability of the miRNA is critical to ensure therapeutic efficacy, and an increase in degradation rate of miRNA-22 causes reduction in %TGI (as revealed by LSA, **Figure 4b**), thereby reinforcing the need for NP-mediated delivery to protect the cargo until delivered to the cytosol. In the second bracker, tumor-specific parameters controlling PD-L1 and eEF2K protein production (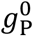 and 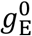, respectively), and the efficiency of PD-L1 (*ε*_P_) at inhibiting immune cell-induced tumor death rank second in GSA, suggesting that PD-L1-mediated tumor immunosurveillance blockade and eEF2K-induced tumor proliferation and PD-L1 production that affect the intrinsic tumor growth and death, are important determinants of miRNA-22 efficacy. This suggests that delivering anti-PD-L1 therapies to target high PD-L1 activity can improve treatment outcomes when used in combination combination with miRNA-22.

These parameters are followed by NP size (*ϕ*_NP_) and NP degradability (*δ*_NP_). NP characteristics strongly influence the systemic pharmacokinetics of NPs (driven by hepatic and renal clearance (42, 60-62)) and also NP transport to and accumulation within the tumor (driven by extravasation across tumor vasculature, diffusion through tumor interstitium, endocytosis into cancer cells, and metabolism-dependent degradation in the cancer cell cytosol (4, 22)). These NP-specific parameters rank highly for their influence on %TGI due to their role in miRNA-22 delivery to the tumor. Of note, as for the individual effects of NP size (*ϕ*_NP_), we observed an inverse monotonic trend between %TGI and the investigated parameter values, suggesting that an increase in NP size leads to reduced %TGI (**Figure 4b**). This suggests that while smaller NPs have smaller drug loading capacity, this may be compensated by using larger quantities to deliver the same dose of drug, which can then outperform larger NPs primarily due to better pharmacokinetics and greater tumor penetration. Note that the corresponding number of NPs injected to deliver 0.026 mg/kg miRNA-22 via NPs of different sizes in our study ranged from ∼90 billion (for size 350 nm) to ∼1.5 quadrillion (for size 14 nm), which lies well within the range of values used in preclinical studies and clinical trials (63). While we did not investigate renally clearable NPs (<10 nm) due to lack of reported clinical application for drug delivery, we anticipate poorer performance from such particles, primarily due to their short circulation half-life driven by rapid renal clearance (42, 64). Further, within the same ranking bracket is the parameter governing the induction of eEF2K degradation by miRNA-22 (*A*_M,E_), suggesting the expected significance of miRNA-22 for eEF2K degradation to inhibit tumor growth.

Parameters in the 4th and 5^th^ ranking brackets include those that control the positive feedback between eEF2K protein and tumor growth (*A*_E,B_, *A*_B,E_), the half-maximal effective concentration of miRNA-22 in suppressing eEF2K 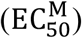, the potency of eEF2K in inducing tumor growth (*K*_E,B_), and importantly, the tumor microvascular surface area (*S*). This suggests that the positive feedback role of eEF2K with tumor growth is secondary relative to its role in immune suppression (these effects are found in the second bracket), and the primairy effective mechanism of action of miRNA-22 is immume suppression, and associate reduction of tumor growth represents a beneficial – but secondary – therapeutic mechanism. The importance of tumor microvascular surface area is attributed to its role in determining rate of extravasation of NPs across tumor vasculature for drug delivery to the cells. However, tumor blood flow rate (*Q*) does not appear to have a significant impact on therapy efficacy.

Together, our simulations find that tumor response is most sensitive to the immune checkpoint effects of PD-L1, miRNA-22-eEF2K interaction, miRNA-22 potency and stability, and NP characteristics. This finding may provide opportunities for patient-specific optimization of NP-mediated miRNA-22 therapy. It also warrants the use of combination therapies, particularly immune checkpoint inhibitors in combination with with chemotherapeutics due to the chemoresistive influence of eEF2K, in order to achieve a better treatment outcome. In light of the ranking obtained through GSA, we understand that LSA may not be required to obtain a ranked order of parameters for their influence on %TGI, because unlike GSA, LSA does not incorporate the interactions between parameters that may influence the outcome, and thus only provides a limited assessment into the sensitivity of parameters. However, LSA can still be used to obtain the empirical relationships between individual parameters and model output, as shown in **Figure 4b**.

### Combination therapies, population variability, and synergy

We then sought to test the effects of combining miRNA-22 with standard-of-care drugs for TNBC, i.e., chemotherapeutics (doxorubicin) and immune checkpoint inhibitors (Atezolizumab) for improvement in %TGI outcome. For these numerical experiments, clinically relevant doses of 2.4 mg/kg Q3W (once every three weeks) doxorubicin and 2 mg/kg Q3W Atezolizumab were simulated for the representative virtual patient (shown in **Figure 3a**) in various combinations with 0.026 mg/kg QW miRNA-22 given for six months, starting 80 weeks after tumor initation. As shown in **Figure 5a**, combining miRNA-22 nanotherapy with the immune checkpoint inhibitor improves the outcome from intermediate response to partial response (30% < TGI ≤ 50%), and combined with doxorubicin it leads to major response (TGI > 50%), which almost reaches complete response when the three modalities are given together. Further, to assess the effects of patient variability and tumor heterogeneity on drug combination outcomes, 2,000 virtual patients were generated as before, and as shown in **Figure 5b**, the three drug combination therapy produced major response in ∼60% of patients, which is three times the patients that showed major response with QW miRNA-22 monotherapy (**Figure 3c**). However, the response worsened when miRNA-22 was given Q2W, either alone or in combination (**Figure 5c**).

**Figure 5.**
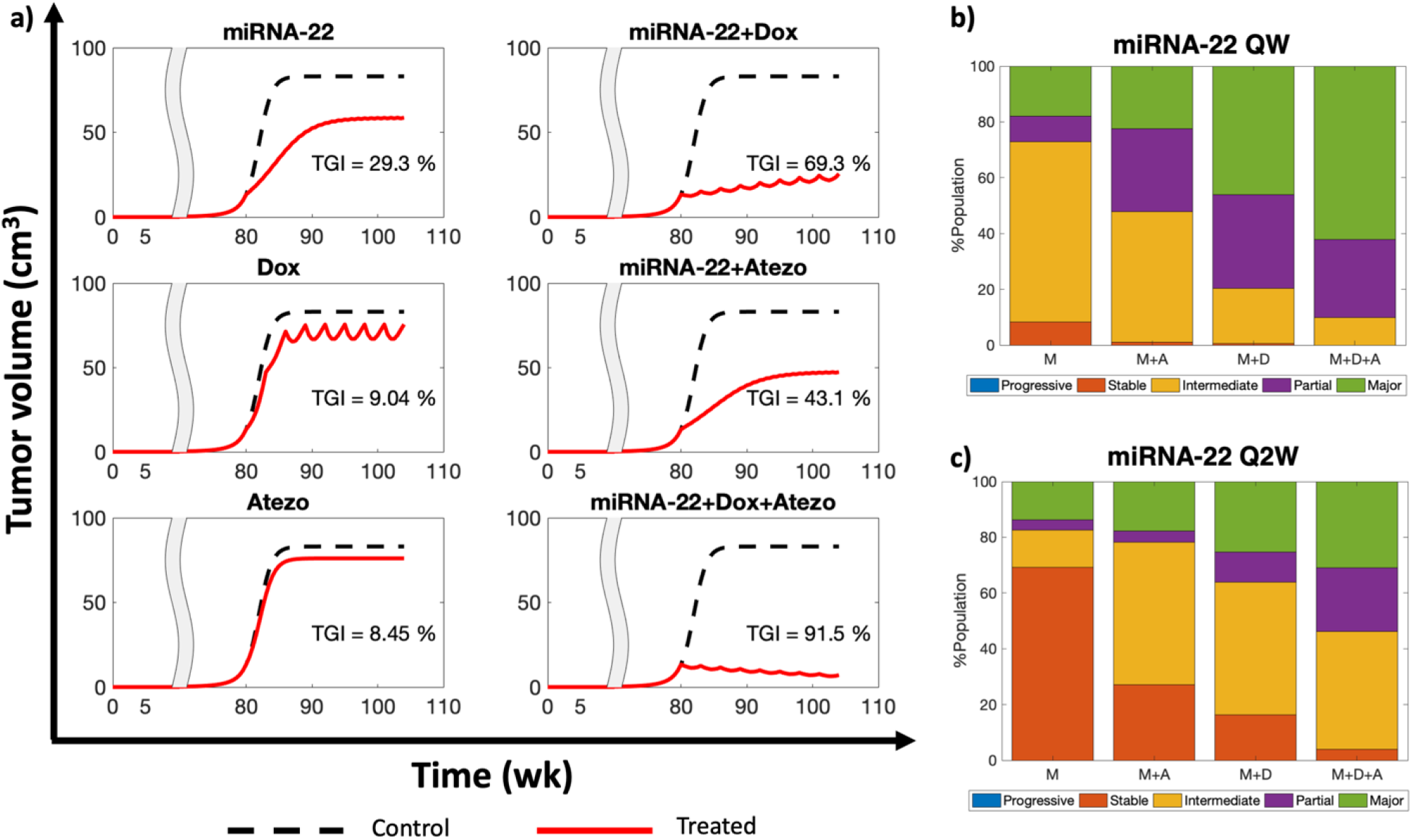
Combination therapies. **a**) Effects of QW dose of miRNA-22 (M) alone or in combination with doxorubicin (Dox or D), or Atezolizumab (Atezo or A) on %TGI are show. **b**,**c**) Effects of inter-individual variability on %TGI following treatment with combination therapies is shown when miRNA-22 is given **b**) QW or **c**) Q2W. Note that the other three drugs were administered once every three weeks (Q3W) in all cases.

Finally, our observation that monotherapies without miRNA-22 show stable disease under the given treatment protocols (**Figure 5a**), while two or three drug combinations with miRNA-22 produce significant improvements in treatment outcome, warrants testing for the occurrence of drug synergy between miRNA-22 and other drugs. For this, as shown in **Figure 6a**, %TGI was calculated through model simulations for various doses of three monotherapies (miRNA-22 QW, doxorubicin Q3W, Atezolizumab Q3W) and three combination therapies (miRNA-22 QW + doxorubicin Q3W, miRNA-22 QW + Atezolizumab Q3W, miRNA-22 QW + doxorubicin Q3W + Atezolizumab Q3W), and was used as an input for Chou-Talalay method (58) to calculate the combination indices (CI) of drug combinations. As shown in **Figure 6b**, CI values <1 for the three combinations of miRNA-22 indicate drug synergy with doxorubicin, Atezolizumab, and doxorubicin+Atezolizumab.

**Figure 6.**
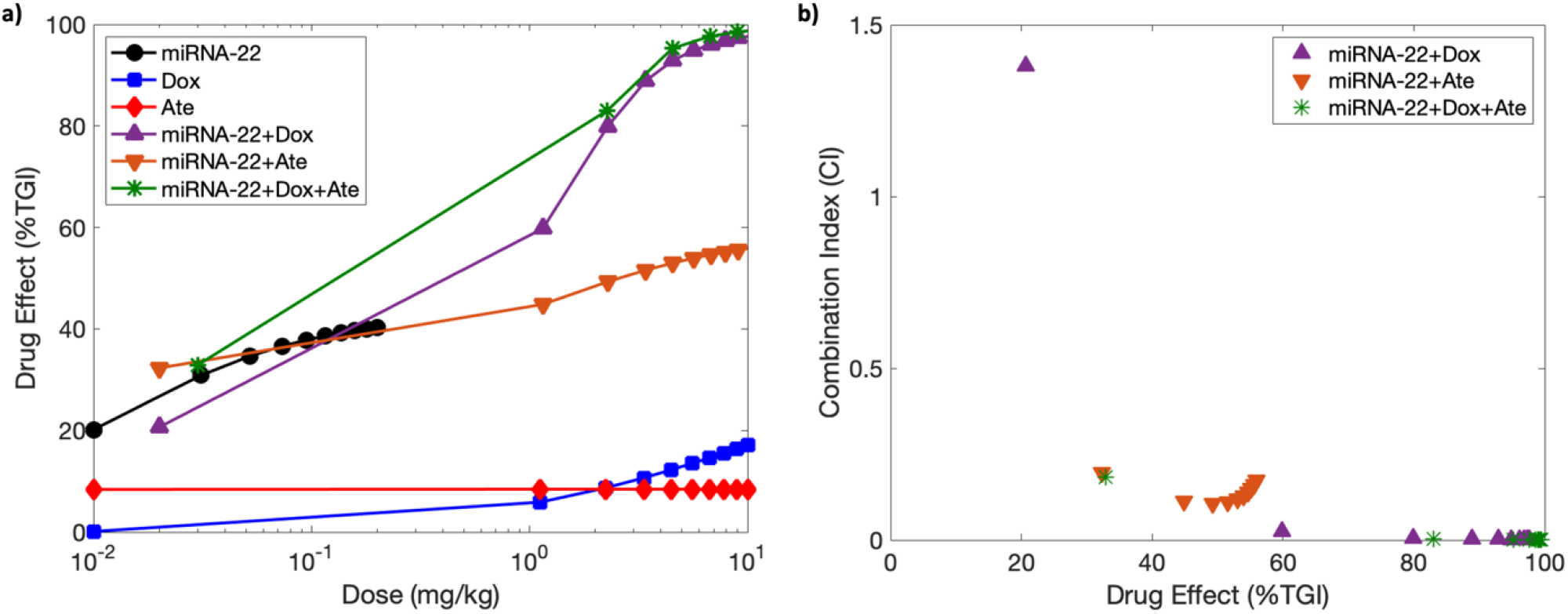
Drug synergism. **a**) Dose-response data generated from model simulations for various monotherapies and combination therapies. Note: Dox indicates doxorubicin and Ate represents Atezolizumab. **b**) Combination indices calculated with the Chou-Talalay method identify drug synergy for various combinations of miRNA-22. CI<1 indicates drug synergism.

## Conclusion

Using a multiscale mechanistic modeling approach, we studied the translational PK-PD of NP-loaded miRNA-22 as a therapeutic for TNBC, alone or in combination with other FDA approved therapeutics. For this, the model was first calibrated with published *in vivo* data involving treatment of MDA-MB-231 tumor-bearing mice with miRNA-22, doxorubicinin, or an immunecheckpoint inhibitor. The calibrated model was extrapolated to the human scale by substituting the physiological parameter values of mice with humans, or by scaling of the parameters with standard allometric techniques. Using the extrapolated model, the dose-response curves and effects of inter-individual variability on treatment outcome were assessed and quantified through a scale analogous to RECIST 1.1. Percent tumor growth inhibition (%TGI) saturated at a dose of 0.05 mg/kg, irrespective of the treatment frequency and doubling time of the tumor. For our translational analysis, a dose of 0.026 mg/kg was used, obtained through allometric scaling of dose for mice. By creating a virtual patient population through perturbation of patient-specific parameters, patient response to variable miRNA doses was quantified, and it was observed that at 0.02 mg/kg, the fraction of patient population showing major response (≥ 50% TGI) to therapy saturated at ∼40%. Within the scope of our computational investigation, further increment in the dose only increased the fraction of partial responders (i.e., patients exhibiting ≥ 30% and < 50% TGI), and appeared to saturate at 0.14 mg/kg with ∼35% patients exhibiting < 30% TGI above that dose.

Parameter sensitivity analysis was conducted to identify the key determinants of %TGI in miRNA-22 nanotherapy. This analysis revealed the significance of miRNA-22-eEF2K interaction, eEF2K-tumor growth feedback loop, tumor vascularization, miRNA-22 potency and stability, NP characteristics, and also the immunecheckpoint effects of PD-L1, which highlights the potential of combination with anti-PD-L1 therapy to improve %TGI. This was supported by numerical experiments involving the combination of NP-loaded miRNA-22 with a clinically used immune checkpoint inhibitor (Atezolizumab) and/or doxorubicin. The triple combination of miRNA-22 with doxorubicin and this immune checkpoint inhibitor lead to almost three-fold increase in population fraction exhibiting major response in comparison to miRNA-22 alone. Importantly, the suspected drug synergy between miRNA-22 and doxorubicin and immunecheckpoint inhibitors was confirmed through the Chou-Talalay combination, whereindices were found to be < 1.

Our analysis, based on a well-calibrated mathematical model extrapolated to the human scale, provides valuable pre-translational, quantitative insights into the limitations, challenges, and opportunities associated with the translation of miRNA-22 nanotherapy for TNBC patients. The ability of the model to explore the effects of patient variability and tumor heterogeneity through parameter perturbation and sampling demonstrates the utility of our in-silico tool to conduct *virtual* clinical trials to assess the effects of anticancer therapeutic agents, which can provide immediate feedback to biologists and clinicians regarding potential problems and their solutions to support the preclinical development and clinical translation of novel therapeutics. The model presented here captures the key processes involved in systemic pharmacokinetics of nanoparticles; however, for a more detailed characterization, we will integrate the tumor compartment with a whole-body physiologically-based pharmacokinetic model in future. Also, spatial tumor heterogeneity, genetic variability, and a more complete tumor microenvironment (with emphasis on immune cells) will be introduced in the tumor compartment of the model to further explore the effects of heterogeneity in drug diffusion barriers, drug resistant cell populations, and tumor immunosurveillance.

## Data Availability

All data produced in the present study are available upon reasonable request to the authors.

## Author Contributions

ZW conceived the study and conceptualized the idea. PD, ZW designed the model and the model analysis. PD, JRR developed the model. PD performed the model analysis. PD, JRR, JDB, MJP, GAC, BO collected the data. PD, JRR, JDB, MJP, CC, AHN, RP, WA, VC, GAC, BO, ZW interpreted the results. PD, JRR, JDB, MJP, CC, AHN, RP, WA, VC, GAC, BO, ZW wrote or edited the manuscript.

## Acknowledgements

The research work was supported by the National Science Foundation grant DMS-1930583 (VC, ZW), National Institutes of Health grants 1R01CA253865 (VC, BO, ZW), 1U01CA196403 (VC, ZW), 1U01CA213759 (VC, BO, ZW), 1R01CA226537 (RP, WA, VC, ZW), 1R01CA222007 (VC, GAC, BO, ZW), U54CA210181 (VC), and the Cockrell Foundation (PD). GAC also acknowledgs the Felix L. Haas Endowed Professorship in Basic Science, and the following grant support: NCI (1R01 CA182905-01, 1R01CA222007-01A1), NIGMS (1R01GM122775-01), DoD (Idea Award W81XWH2110030), a Team DOD grant in Gastric Cancer, a Chronic Lymphocytic Leukemia Moonshot Flagship project, a CLL Global Research Foundation 2019 grant, a CLL Global Research Foundation 2020 grant, a Mathers Foundation grant, an Institutional Research and Development Grant associated with the Brain SPORE 2P50CA127001. The funders had no role in study design, data collection and analysis, decision to publish, or preparation of the manuscript.

