## Supplementary material for "Translational Modeling Identifies Synergy between Nanoparticle-Delivered miRNA-22 and Standard-of-Care Drugs in Triple Negative Breast Cancer": SI

***Short title:*** Modeling miR-22 therapy in TNBC
**Keywords:** Cancer treatment, mathematical modeling, microRNA, pharmacokinetics and pharmacodynamics, precision medicine, tumor-immune interaction, allometry

### Invited Contribution

Correspondence should be addressed to:

**Zhihui Wang, Ph.D.**
Associate Professor, Mathematics in Medicine Program
The Houston Methodist Research Institute


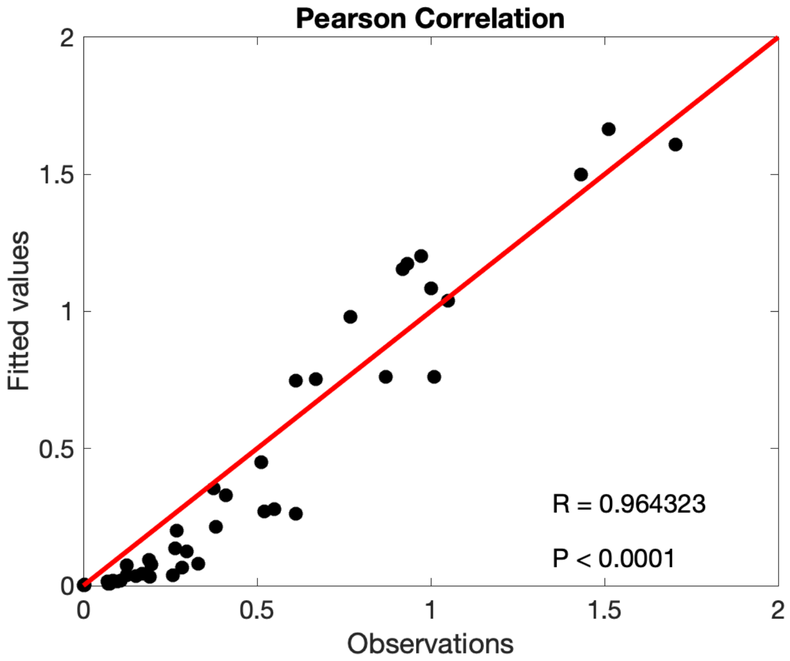


**Figure S1.** Pearson correlation of model fits to *in vivo* data shown in Figure 2.

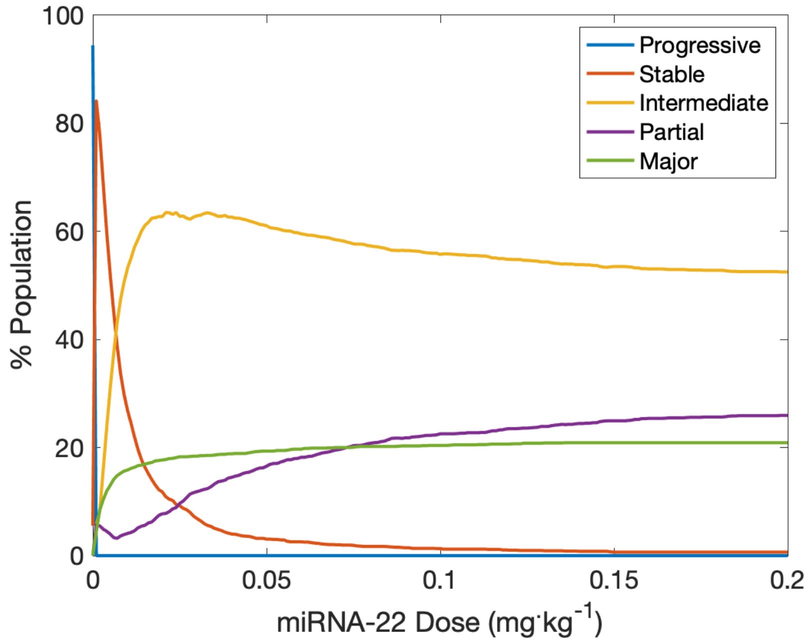


**Figure S2.** Population-scale dose response relationship of miRNA-22 therapy.

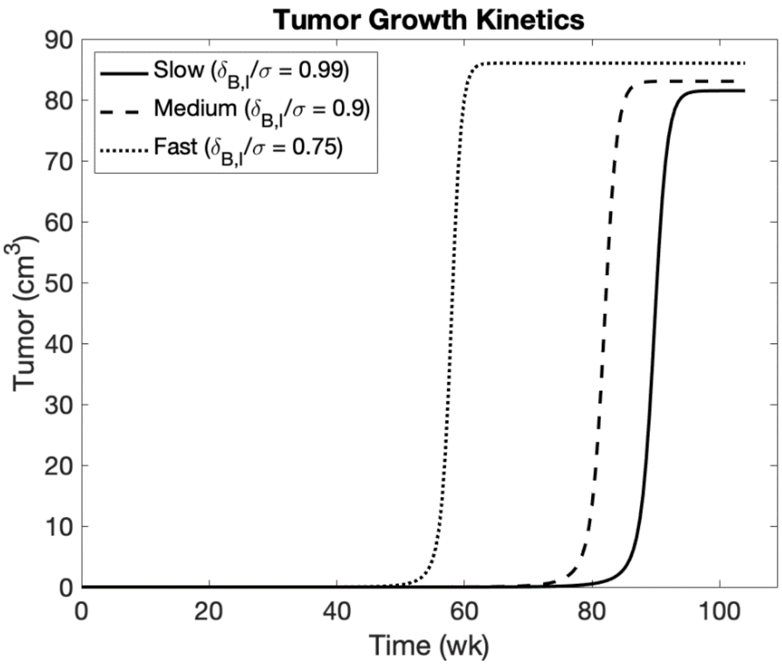


**Figure S3.** Tumor growth kinetics of slow-, medium-, and fast-growing tumors under the control conditions.

### CompuSyn Report

**Experiment Name:** AAPS paper
**Date:** 7/14/21
**File Name:** C:\Users\prash\Desktop\report.cse
**Description**

**Drug:** mirna22 (mirna) [mg/kg]
**Drug:** dox (dox) [mg/kg]
**Drug:** ate (ate) [mg/kg]
**Drug Combo:** mirna+dox (mirdox) (mirna+dox)
**Drug Combo:** mirna+ate (mirate) (mirna+ate)
**Drug Combo:** mirnadoxate (mda) (mirna+dox+ate)

---

Data for Drug: mirna [mg/kg]

| Dose | Effect |
|---|---|
| 0.01 | 0.2017 |
| 0.0311 | 0.3087 |
| 0.0522 | 0.3466 |
| 0.0733 | 0.3662 |
| 0.0944 | 0.3782 |
| 0.1156 | 0.3864 |
| 0.1367 | 0.3922 |
| 0.1578 | 0.3966 |
| 0.1789 | 0.4001 |
| 0.2 | 0.4029 |

10 data points entered.
**X-int:** -0.2695
**Y-int:** 0.08429 +/- 0.03407
**m:** 0.31277 +/- 0.02925
**Dm:** 0.53764
**r:** 0.96676

---

Data for Drug: dox [mg/kg]

| Dose | Effect |
|---|---|
| 0.01 | 0.0012 |
| 1.12 | 0.0589 |
| 2.23 | 0.087 |
| 3.34 | 0.1068 |
| 4.45 | 0.1225 |
| 5.56 | 0.1352 |
| 6.67 | 0.146 |
| 7.78 | 0.1553 |
| 8.89 | 0.1635 |

| Dose | Effect |
| --- | --- |
| 10.0 | 0.1708 |

10 data points entered.

**X-int:** 1.81569
**Y-int:** -1.3608 +/- 0.02420
**m:** 0.74946 +/- 0.02588
**Dm:** 65.4173
**r:** 0.99526

---

Data for Drug: ate [mg/kg]

| Dose | Effect |
| --- | --- |
| 0.01 | 0.08402 |
| 1.12 | 0.08454 |
| 2.23 | 0.08454 |
| 3.34 | 0.08455 |
| 4.45 | 0.08455 |
| 5.56 | 0.08455 |
| 6.67 | 0.08455 |
| 7.78 | 0.08455 |
| 8.89 | 0.08455 |
| 10.0 | 0.08455 |

10 data points entered.

**X-int:** 1033.99
**Y-int:** -1.0352 +/- 1.13E-4
**m:** 0.00100 +/- 1.20E-4
**Dm:** Infinit
**r:** 0.94670

---

Data for Non-Constant Combo: mirdox (mirna+dox)

| Dose mirna | Dose dox | Effect |
| --- | --- | --- |
| 0.01 | 0.01 | 0.2064 |
| 0.0311 | 1.12 | 0.5982 |
| 0.0522 | 2.23 | 0.7995 |
| 0.0733 | 3.34 | 0.8892 |
| 0.0944 | 4.45 | 0.9291 |
| 0.1156 | 5.56 | 0.9493 |
| 0.1367 | 6.67 | 0.9609 |
| 0.1578 | 7.78 | 0.9683 |
| 0.1789 | 8.89 | 0.9733 |
| 0.2 | 10.0 | 0.9769 |

10 data points entered.

---

Data for Non-Constant Combo: mirate (mirna+ate)

| Dose mirna | Dose ate | Effect |
| --- | --- | --- |
| 0.01 | 0.01 | 0.32318 |

| Dose mirna | Dose ate | Effect |
|---|---|---|
| 0.0311 | 1.12 | 0.44847 |
| 0.0522 | 2.23 | 0.49304 |
| 0.0733 | 3.34 | 0.51610 |
| 0.0944 | 4.45 | 0.53018 |
| 0.1156 | 5.56 | 0.53967 |
| 0.1367 | 6.67 | 0.54648 |
| 0.1578 | 7.78 | 0.55162 |
| 0.1789 | 8.89 | 0.55563 |
| 0.2 | 10.0 | 0.55885 |

10 data points entered.

---

Data for Non-Constant Combo: mda (mirna+dox+ate)

| Dose mirna | Dose dox | Dose ate | Effect |
|---|---|---|---|
| 0.01 | 0.01 | 0.01 | 0.32890 |
| 0.0311 | 1.12 | 1.12 | 0.83052 |
| 0.0522 | 2.23 | 2.23 | 0.95271 |
| 0.0733 | 3.34 | 3.34 | 0.97687 |
| 0.0944 | 4.45 | 4.45 | 0.98529 |
| 0.1156 | 5.56 | 5.56 | 0.98933 |
| 0.1367 | 6.67 | 6.67 | 0.99164 |
| 0.1578 | 7.78 | 7.78 | 0.99312 |
| 0.1789 | 8.89 | 8.89 | 0.99414 |
| 0.2 | 10.0 | 10.0 | 0.99488 |

10 data points entered.

---

Median-Effect Plot for Drugs

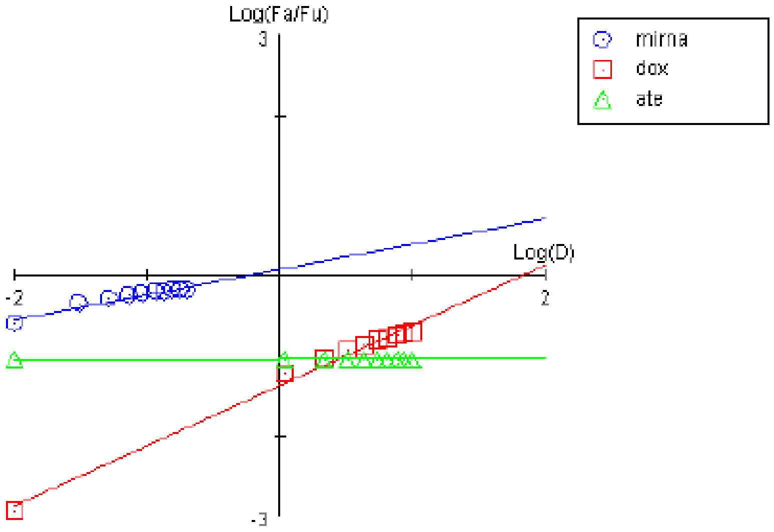

Median-Effect Plot for Drug Combos

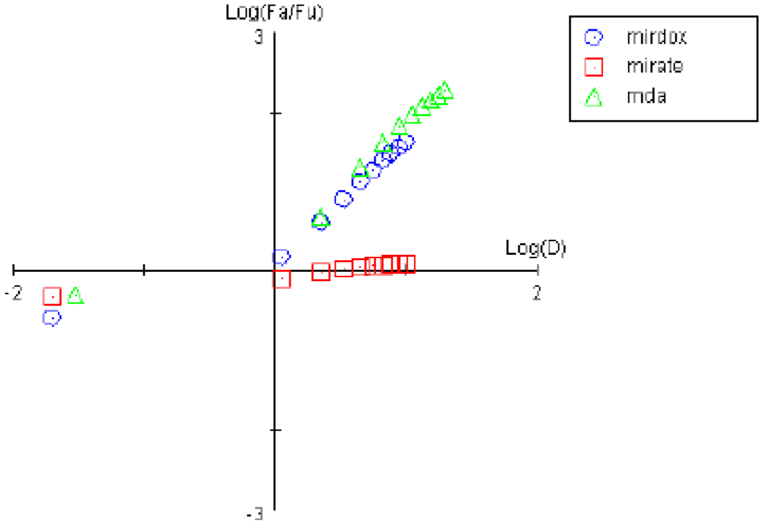


CI Data for Non-Constant Combo: mirdox (mirna+dox)

| Dose mirna | Dose dox | Effect | CI |
|---|---|---|---|
| 0.01 | 0.01 | 0.2064 | 1.37989 |
| 0.0311 | 1.12 | 0.5982 | 0.02627 |
| 0.0522 | 2.23 | 0.7995 | 0.00655 |
| 0.0733 | 3.34 | 0.8892 | 0.00335 |
| 0.0944 | 4.45 | 0.9291 | 0.00224 |
| 0.1156 | 5.56 | 0.9493 | 0.00172 |
| 0.1367 | 6.67 | 0.9609 | 0.00143 |
| 0.1578 | 7.78 | 0.9683 | 0.00125 |
| 0.1789 | 8.89 | 0.9733 | 0.00112 |
| 0.2 | 10.0 | 0.9769 | 0.00104 |

CI Data for Non-Constant Combo: mirate (mirna+ate)

| Dose mirna | Dose ate | Effect | CI |
|---|---|---|---|
| 0.01 | 0.01 | 0.32318 | 0.19766 |
| 0.0311 | 1.12 | 0.44847 | 0.11207 |
| 0.0522 | 2.23 | 0.49304 | 0.10613 |
| 0.0733 | 3.34 | 0.51610 | 0.11095 |
| 0.0944 | 4.45 | 0.53018 | 0.11930 |
| 0.1156 | 5.56 | 0.53967 | 0.12933 |
| 0.1367 | 6.67 | 0.54648 | 0.14007 |
| 0.1578 | 7.78 | 0.55162 | 0.15131 |
| 0.1789 | 8.89 | 0.55563 | 0.16287 |
| 0.2 | 10.0 | 0.55885 | 0.17465 |

CI Data for Non-Constant Combo: mda (mirna+dox+ate)

| Dose mirna | Dose dox | Dose ate | Effect | CI |
|---|---|---|---|---|
| 0.01 | 0.01 | 0.01 | 0.32890 | 0.18226 |
| 0.0311 | 1.12 | 1.12 | 0.83052 | 0.00241 |
| 0.0522 | 2.23 | 2.23 | 0.95271 | 6.27E-4 |
| 0.0733 | 3.34 | 3.34 | 0.97687 | 3.47E-4 |
| 0.0944 | 4.45 | 4.45 | 0.98529 | 2.49E-4 |
| 0.1156 | 5.56 | 5.56 | 0.98933 | 2.02E-4 |
| 0.1367 | 6.67 | 6.67 | 0.99164 | 1.74E-4 |
| 0.1578 | 7.78 | 7.78 | 0.99312 | 1.56E-4 |
| 0.1789 | 8.89 | 8.89 | 0.99414 | 1.44E-4 |
| 0.2 | 10.0 | 10.0 | 0.99488 | 1.35E-4 |

Combination Index Plot

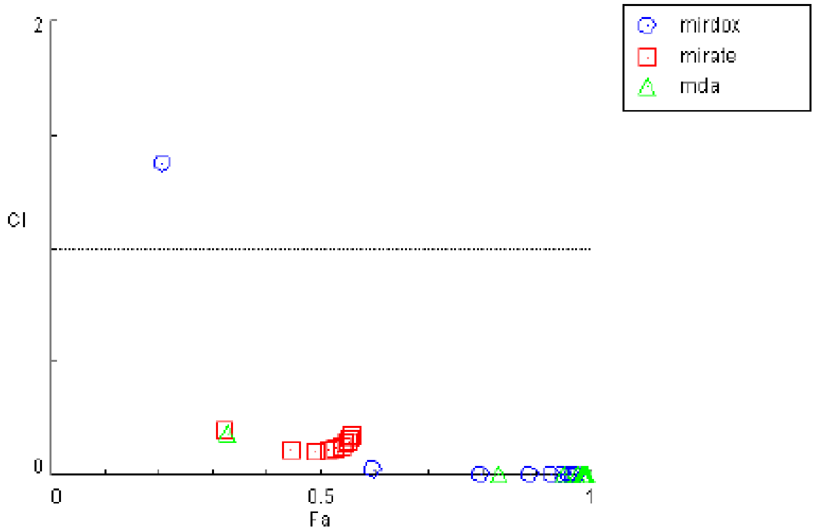
